# Perceptions of Minority Ethnic Groups on Mental Health Electronic Health Records: A Study Protocol

**DOI:** 10.64898/2026.02.09.26345920

**Authors:** Ammarah Ikram, Sahdia Parveen, Dianne Wepa, Clare McGuinn, Eleftheria Vaportzis

## Abstract

Electronic Health Records (EHRs) have not been widely implemented in mental health settings, representing a significant gap in digital health care transformation. A reason for underutilisation includes concerns from healthcare professionals regarding the collection and storage of patients’ sensitive information. Language use can positively influence clinician-patient relationships, and stigmatising language in EHRs viewed by patients could undermine trust. This is concerning as using EHRs have benefits which allow patients to feel safe and empowered regarding their care. Moreover, minority ethnic groups have been found to disengage with EHRs and are more likely to access mental health services through crisis pathways. This qualitative study in collaboration with Bradford District Care NHS Foundation Trust comprises two stages to explore minority ethnic perspectives on mental health EHRs and develop recommendations for their implementation. Stage one investigates minority ethnic service user’s perceptions on EHRs and explores mental health professional’s understanding regarding the sharing of EHRs with service users from minority ethnic groups. The workshops in stage two will use an Experience-Based Co-Design approach to produce practical recommendations for EHR implementation in mental health settings. Participants include minority ethnic service users, mental health professionals, stakeholders, and relevant bodies such as mental health organisations and Information Technology experts utilising EHRs. Data will be gathered through semi-structured interviews, focus groups and workshops, and analysed using reflexive thematic analysis. The study was approved under the Integrated Research Application System (IRAS ID: 348764) and Health Research Authority and Health and Care Research Wales. Findings will be disseminated via social media, blogs, conferences, journals, academic articles, and community and staff meetings held by the Trust. An executive summary will be shared with participants who consented to receive the results.

## Introduction

Within the United Kingdom (UK) challenging the disparities individuals may encounter when accessing mental healthcare services is a significant policy issue (1). Minority ethnic groups are more likely to face adverse encounters from psychological services (2), and struggle with poorer outcomes regarding their mental healthcare (1). For example, clinicians may face complications in providing high levels of care because of cultural and linguistic obstacles (3), which in turn may prevent minority ethnic groups from accessing healthcare services. Minority ethnic groups were more likely to be detained under the Mental Health Act (4). As a result, minority ethnic groups may use mental healthcare services less regularly due to the clinicians’ limited understanding of the cultural competency (1).

EHRs have become an integral part of modern healthcare and represent a development from paper based medical records to more portable and interactive tools such as web-based patient portals and mobile apps (5, 6). Systems such as Open Notes which is a feature or practice within the EHR can provide patients online access to their own clinical documentation (7). Within mental health settings, the implementation of EHRs is relatively slow compared with other health contexts (8). Statistics from the United States (US) showed that compared to general medicine and surgical hospitals that provide access to EHRs, only 49% of psychiatric hospitals have certified EHRs as of 2017 (8). This number was lower than rehabilitation (89%), children’s (87%), and acute long-term care (59%) hospitals (8). However, as these statistics are from the US, it is difficult to generalise these findings to the UK. Because EHRs are required to hold sensitive data, incomplete records may result in patients having to re-live traumatic events due to their data not being fully documented within EHRs (9). The adoption of EHRs in the context of mental health is minimal as consistent detailed mental health documentation cannot be simplified to the fields within EHRs (10). Research highlights mental healthcare professionals have a low willingness to include confidential and sensitive information in EHRs and preferred to limit EHR access to patients (11). Findings from a scoping review revealed service users became distressed when viewing their EHRs due to inaccurate notes and perceived disrespectful language which adversely affected the service users’ experiences when accessing mental health care (12). EHRs can create barriers specifically for minority ethnic groups as these groups may have concerns regarding their mental health treatments and negative experiences with health providers (1).

The aim of the study is to understand minority ethnic groups perspectives on mental health EHRs. The study will address the following research questions: (1) how are electronic mental health records perceived and used by people from minority ethnic groups; and (2) what are mental health professionals’ concerns regarding the sharing of EHRs with service users from minority ethnic groups; and (3) what recommendations can be made to support the implementation of EHRs in mental health settings from service users from diverse ethnic backgrounds. The study’s objective outputs include (1) to examine how open, shared mental health, EHRs are perceived by different minority ethnic groups and probe their potential level of engagement; (2) to investigate the participants’ perceptions of the utility of open access EHRs; (3) determine participants’ perceived barriers and facilitators to access their open shared EHRs; (4) explore mental health professionals’ barriers and facilitators regarding sharing the EHRs with service users from minority ethnic groups; and (5) produce evidence-based recommendations for implementation of EHR access across different minority ethnic groups.

## Material and Methods

The proposed study will address the aims and research questions using a qualitative approach which is necessary to understand individual experiences (13). This approach will also allow the development of concepts that clarify the phenomena of EHRs in natural, rather than experimental settings (14). Emphasis is given to the meanings, views and experiences of all the participants being researched (14). In stage one, the first research question (i.e., how are electronic mental health records perceived and used by minority ethnic groups) will be investigated through semi-structured interviews with minority ethnic service users accessing mental healthcare. With the nature of the research topic being sensitive, one to one semi-structured interviews are a suitable method for minority ethnic service users rather than focus groups. The second research question (i.e., what are mental health professionals’ concerns regarding the sharing of EHRs with service users from minority ethnic groups) will be explored through focus groups with mental health professionals. Focus groups were used in this study to explore professionals’ attitudes, beliefs, and experiences as the interactive format allows participants to build on each other’s contributions (15). This method is suitable for identifying barriers and facilitators to using EHRs in mental healthcare as it allows participants to discuss and reflect on shared experiences in a group setting.

In stage two an Experience Based Co-Design (EBCD) approach will be adopted to address the third research question (i.e., what recommendations can be made to support the implementation of EHRs in mental health settings from service users from diverse ethnic backgrounds). EBCD is an approach enabling staff and service users to co-design services together in partnership (16). EBCD places the patients’ experiences at the centre of the design process (17). The findings from the interviews and focus groups conducted in stage one of the study will be presented during the co-design workshops in stage two to inform the development of recommendations for implementing EHRs in mental health settings within UK mental health trusts. Stakeholders and professional bodies with experience of EHRs, alongside mental health professionals and service users accessing mental healthcare, will participate in the workshops to collaboratively prioritise key issues and co-develop practical solutions. The outputs of these workshops will inform service improvements, evidence-based recommendations, and implementation strategies aimed at supporting equitable and effective access to EHRs for service users.

For stage one recruitment will begin Monday 23^rd^ June 2025 and end Friday 12^th^ December. For stage two recruitment will begin Monday 2^nd^ March 2026 and end Friday 29^th^ May 2026.

### Patient Public Involvement

The Patient and Public Involvement (PPI) coordinator lead from the University of Bradford appointed a PPI group for the study. The PPI group consist of two members with lived experience of mental health services, both from minority ethnic backgrounds. The group meets every three months to provide input and feedback on study materials, procedures, and interpretation of findings. More specifically, the PPI group will ensure the materials such as interview, focus group and workshop guides are suitable guaranteeing the diverse needs of the participants are supported throughout the research. The PPI group will co-validate the themes found during the data analysis. The PPI group will be presented short, anonymised sections of the data and receive feedback regarding interpretation. The PPI group will also be involved in producing and disseminating user-friendly accessible updates and summaries of the findings of the study. The impact of the PPI members involvement will be recorded through meeting notes, reflective logs of implemented changes, and direct feedback on how their input shaped study decisions and outputs.

### Sample

A purposive stratified sampling approach will be used to recruit participants. Recruitment will take place through social media platforms, mental health organisations, and organisationally through the Bradford District Care NHS Foundation Trust (BDCFT), a collaborator of this study. The study will aim to recruit a total of approximately 50 participants across the two stages. In stage one, around 30 participants will be recruited for interviews and focus groups to explore perspectives on electronic health records (EHRs). This will include approximately 9–15 individual interviews and 2–3 focus groups, each comprising 5–6 participants. These numbers are informed by research from Hennink and Kaiser (18), who found that data from 9–17 interviews are typically sufficient to achieve data saturation, and by studies suggesting that 2–3 focus groups capture around 80% of the most prevalent themes.

Participants from stage one will be invited to take part in stage two to ensure continuity. Stage two will involve approximately 20 participants in two workshops. The first workshop will include 5–8 minority ethnic service users accessing mental health services, and the second will include 5–8 mental health professionals, stakeholders, and relevant bodies. If recruitment for either stage falls short, additional recruitment and advertising will be conducted until sufficient data saturation is achieved.

### Population and Recruitment

The population of interest includes minority ethnic service users accessing mental health care, mental health professionals, stakeholders and relevant bodies such as mental health organisations and IT experts utilising EHRs. The research will be carried out within the District of Bradford where the Asian population increased from 26.8% in 2011 to 32.1% in 2021 and is the second highest minority ethnic group in the city after the White population (19). It is expected that individuals from Asian, Black, Mixed and White other groups accessing mental health services will be recruited. The underrepresentation of these communities in both primary care services such as mental healthcare (1), and in health and social research (20) highlights the need in determining the perceptions of EHRs from minority ethnic groups.

In stage one, the study will recruit minority ethnic service users and mental health professionals from secondary, tertiary, community mental health services and inpatient settings across BDCFT, as well as from mental health organisations. Recruitment will be supported by BDCFT through internal communications, staff meetings, posters in community hubs and waiting areas, and local research databases. Social media platforms such as Facebook, Instagram, and X will be used to recruit participants. Participants from stage one will be invited to take part for continuity in stage two. Additional recruitment may include new service users, professionals, stakeholders, mental health organisations, charities, and IT experts using EHRs. Recruitment will reflect stage one methods in stage two. For stage one and stage two recruitment materials may be translated into Urdu if required to support inclusion. This is because Urdu is the third language spoken within Bradford and the first author (AI) is a proficient speaker of Urdu allowing recruitment to not be limited to individuals speaking English and offers the opportunity for inclusion as widely as possible across the population of Bradford (21).

Interested participants will be contacted by the research team through their preferred methods, phone, email, social media. Participants meeting the eligibility criteria of the research will be provided a participant information sheet and consent form. A 1-week period is provided allowing participants to reflect, ask any questions and promotes free choice. The process will follow the UK Mental Capacity Act 2005 and participants can withdraw at any point up to two weeks post participation. The recruitment process will remain accessible, voluntary, and sensitive. Further recruitment will be carried outside the Bradford district if an adequate number of participants is not recruited

### Eligibility Criteria

#### Inclusion Criteria

Participants eligible for this study must be over the age of 18 and of any gender. Participants must be from a minority ethnic group and are past or present service users accessing mental healthcare, mental health professionals, and for stage two stakeholders and relevant bodies such as mental health charities, organisations, or IT experts utilising EHRs. To promote inclusivity within the diverse Bradford population, participants must speak either English or Urdu.

#### Exclusion Criteria

Participants will be excluded for practical, ethical, or methodological reasons. Individuals who are unable to provide informed consent for example, due to cognitive impairment or are experiencing an acute mental health crisis are not eligible for this study to avoid causing further distress. Those under the age of 18 are also excluded.

To ensure the study focused on relevant experiences, individuals who have not accessed mental health services or did not have professional, stakeholder, or lived experience related to the research topic are excluded. As the study aimed to explore perspectives within minority ethnic groups, individuals who do not identify as belonging to a minority ethnic background will not be eligible to participate.

### Procedure

Stage one minority ethnic service users meeting the eligibility criteria will take part in One-to-One semi-structured interviews and mental health professionals will participate in focus groups. Participants will receive participant information sheets and consent form prior to participation. Interviews and focus groups are expected to last no longer than two hours. At the end of each interview and focus group, participants will be thanked, debriefed, and informed on how they can receive the study’s findings once the project is completed and a debrief sheet will be provided (see supporting information).

Separate workshops for minority ethnic service users and for mental health professionals, stakeholders, and relevant bodies such as mental health organisations, charities, and IT experts who use EHRs will be conducted. The findings from stage one of the research will be presented in the workshops to discuss the themes which arise and produce recommendations to improve the usability of EHRs in mental health care for minority ethnic service users. Participants will receive participant information sheets and consent sheets. The workshops are expected to last no more than two hours. Participants will be debriefed, thanked for their time and informed on how they can receive the study’s findings once the project is completed. A debrief sheet will be provided.

A blended approach will be used for the interviews, focus groups and workshops and depending on the preferences of the participants these may take place in-person or online via Microsoft Teams. In-person data collection will take place across multiple premises including the University of Bradford, BDCFT and mental health organisations.

### Analysis

In stage one, data from semi-structured interviews and focus groups will be transcribed and entered into NVivo software for qualitative analysis using reflexive thematic analysis (RTA) (22). RTA allows a flexible yet systematic approach to the traditional thematic analysis and values the researcher’s subjectivity in interpreting the data (23). RTA is the appropriate data analysis method as participants lived experiences and perceptions of a particular phenomenon (24), such as EHRs in mental health can be determined. This is necessary in identifying recommendations to encourage EHR implementation in mental health settings for minority ethnic service users. In stage two, data generated during the co-design workshops will also be analysed using RTA to synthesise co-designed insights into key barriers, facilitators, and recommendations for implementation. Together, these analytic processes align with EBCD, which integrates participants experiences with collaborative solution development to inform service improvement.

### Ethics and dissemination

Ethical approval for this study was obtained from Wales Research Ethics Committee 7. Health Research Authority and Health and Care Research Wales Approval was issued to conduct the study. The study was approved under the Integrated Research Application System (IRAS), IRAS ID: 348764.

### Consent and Capacity

Consent will be obtained for both stages of the study at least 1-week before the interviews, focus groups, and workshops. Participants will be required to return consent forms either online by signing electronically or in person in writing within this 1-week period. This period gives time for participants to reflect and ask any questions and promotes free choice. The research team will then make contact again either by email or telephone, according to the participant’s preferred method of contact and assess capacity to consent to take part, in line with the UK Mental Capacity Act 2005. Capacity will be assessed based on understanding the research purpose, retaining information to decide, and communicating that decision. This ensures participants are engaged and can make informed, voluntary choices.

### Distress and Safeguarding

The nature of the topic may cause distress for minority ethnic service users due to stigma associated with mental health. To address this possibility, participants will be reminded before the data collection their responses will be anonymised, and signposting of relevant bodies will be provided via debrief sheets. The researchers, will monitor for distress, offer breaks or stop sessions if needed and aim to end sessions on a positive note. During each interview, focus group and workshop, a mental health professional from the BDCFT will be available to provide professional support for any participant in distress. Participants will be provided with safe spaces either a separate room in person or a private breakout room online where they can take breaks or access support if they feel distressed during the study. If a safeguarding risk arises, this will be discussed within the research team. If a safeguarding risk is required to be reported the research team will contact the Local Authority Adult Safeguarding Team and act in line with their safeguarding policies and procedures.

### Data Management and Confidentiality

All data will be managed securely and held following the Data Protection Act 2018 and General Data Protection Regulation to ensure the confidentiality and security of the data. Data will be stored securely on the University of Bradford’s password-protected OneDrive. Any paper copies of forms will be uploaded onto the University of Bradford’s password-protected OneDrive and shredded. Audio recordings will be transcribed, pseudonymised, and stored securely. Identifiable information will be limited, and the research team will only have access to this. Participants will be informed that confidentiality cannot be fully guaranteed in group settings. Consent will be requested from participants to use anonymised quotes in reports, publications, and presentations.

### Right to withdraw

After participation all participants will be made aware they have up to two weeks to withdraw their data from the study and after this cut off point their data will be kept and remain anonymised.

## Discussion

The purpose of the study is to examine minority ethnic groups perceptions of open-access EHRs in mental health settings. The study will begin to produce evidence-based recommendations for the implementation of the EHR systems. Within the context of mental health, EHR access is low, and healthcare providers present concerns of including confidential and sensitive medical information, and thus, prefer to limit access to service users. This is paradoxical as EHRs are valuable in improving patient safety, reducing errors, and increasing accessibility and information sharing between clinicians and service users. However, despite these benefits and the low acceptance of electronic systems in mental health settings, research within this domain is limited. Supporting this notion, is a systematic review which is published and was completed by the research team (AI, SP, EV) which aimed to explore, ‘How are electronic mental health records perceived and used by different ethnically diverse groups and what are the barriers and facilitators?’ (25). The review identified eight papers originating from the United States which demonstrates the need for research in the UK. Furthermore, minority ethnic groups are still underrepresented within primary care services such as mental healthcare and in health and social research. Hence, addressing inequalities individuals face when accessing mental healthcare is a policy concern as minority ethnic groups are likely to have adverse experiences from psychological services. Therefore, with such limited research, the purpose of the study is necessary in recognising whether EHRs can be effectively implemented in mental health settings and utilised by minority ethnic groups.

The findings of this research will be disseminated with the aim of improving mental health services, informing policy, and empowering minority ethnic service users. Dissemination will be tailored to different audiences to ensure accessibility and impact. For academics, findings will be included in the PhD thesis, submitted to peer-reviewed journals, and presented at national and international conferences to share insights and study outcomes. For professionals, stakeholders, and policymakers, results will be presented at community and staff meetings held by the BDCFT and shared through executive summaries highlighting key findings to guide service development and improve practice. For service users and the wider public, participants who consent to receive the findings will be provided with plain language executive summaries. Accessible formats such as blogs and social media posts will be used to raise awareness, share insights, and promote engagement with mental health issues. Reasons for this include social media being able to provide an informal and rapid means of notifying people about research by creating links to papers online (26). This ensures that the research is both academically rigorous and practically useful, fostering meaningful impact for all relevant audiences.

## Data Availability

No datasets were generated or analysed during the current study. All relevant data from this study will be made available upon study completion.

## Authors contributions

AI: Methodology, Writing – Original Draft. SP: Conceptualization, Methodology Writing – Reviewing and Editing, Supervision, Funding acquisition. DW: Writing – Reviewing and Editing, Supervision. CM: Conceptualization Writing – Reviewing and Editing, Funding acquisition. EV: Conceptualization, Methodology, Writing – Reviewing and Editing, Project administration, Supervision, Funding acquisition.

